# COVID-19 RELATED IMMUNIZATION DISRUPTIONS IN RAJASTHAN, INDIA: A RETROSPECTIVE OBSERVATIONAL STUDY

**DOI:** 10.1101/2020.12.04.20244327

**Authors:** Radhika Jain, Ambika Chopra, Camille Falézan, Mustufa Patel, Pascaline Dupas

**Affiliations:** Stanford University; Jameel Poverty Action Lab (J-PAL) South Asia; Stanford University, Center for Economic and Policy Research (CEPR), National Bureau of Economic Research (NBER), Jameel Poverty Action Lab (J-PAL)

## Abstract

**Introduction:** Governments around the world suspended immunization outreach to control COVID-19 spread. Many have since resumed services with an emphasis on catch-up vaccinations to reach children with missed vaccinations. This paper evaluated immunization disruptions during India’s March-May 2020 lockdown and the extent to which subsequent catch-up efforts reversed them in Rajasthan, India.

**Methods:** In this retrospective observational study, we conducted phone surveys to collect immunization details for 2,144 children that turned one year old between January and October 2020. We used logistic regressions to compare differences in immunization timeliness and completed first-year immunization status among children that were due immunizations just before (unexposed), during (heavily exposed), and after (post-exposure) the lockdown.

**Results:** Relative to unexposed children, heavily exposed children were significantly less likely to be immunized at or before 9 months (OR 0.550; 95%CI 0.367-0.824; p=0.004), but more likely to be immunized at 10-12 months (OR 1.761; 95%CI 1.196-2.591; p=0.004). They were also less likely to have completed their key first-year immunizations (OR 0.624; 95%CI 0.478-0.816; p=0.001) by the time of survey. In contrast, post-exposure children showed no difference in timeliness or completed first-year immunizations relative to unexposed children, and their immunization coverage was 6.9pp above heavily exposed children despite their younger age. Declines in immunization coverage were larger among children in households that were poorer, less educated, lower caste, and residing in COVID red zones, although subgroup comparisons were not statistically significant.

**Conclusion:** Disruptions to immunization services resulted in children missing immunization during the lockdown, but catch-up efforts after it was eased ensured many children were reached at later ages. Nevertheless, catch-up was incomplete and children due their immunizations during the lockdown remained less likely to be fully immunized 4-5 months after it lifted, even as younger cohorts due immunizations in June or later returned to pre-lockdown schedules.

## INTRODUCTION

The COVID-19 pandemic has caused severe disruptions to health services worldwide, as health system resources have been reallocated to the pandemic response, supply chains have been disrupted, and travel barriers and fear of contracting the virus have deterred health care utilization.[1–3] Immunization services are among the key preventive health services that have been affected in both rich and lower income countries.[4–6]

In late March 2020, the WHO issued guidelines to countries to prioritize immunization as a core health service, but to temporarily suspend mass vaccination campaigns and weigh local risks and benefits in decisions on whether to continue immunization outreach activities.[7] More than half of the 129 countries covered in a survey released in May reported complete suspensions or substantial disruptions to immunization services in March and April, putting an estimated 80 million children under 1 year old at increased risk of contracting vaccine preventable diseases.[8] Travel and access barriers, supply-side disruptions due to provider shortages and suspended immunization services, and user fears of contracting COVID-19 have emerged as key reasons for reductions in immunizations.[8,9]

Disruptions to routine and critical care during large disease outbreaks can have sizeable effects on health outcomes. Sharp decreases in health care utilization during the Ebola and SARS epidemics were estimated to have indirect mortality effects similar in magnitude to the direct effects of the virus.[10–14] Declines in routine immunizations led to significant outbreaks of vaccine preventable diseases like Measles.[15–17] Although young age and female sex are associated with lower COVID-19 mortality risk, early prediction models suggest that the indirect mortality effects of the pandemic among women and children due to lower use of preventive services may be substantial.[18,19] A modeling analysis covering 54 African countries found that sustaining routine child immunizations would prevent substantially more deaths than excess COVID-19 mortality risk from vaccination clinic visits would cause.[20]

Studies have documented large decreases in immunization coverage in higher income countries during the early months of the pandemic, when countries had imposed social distancing and quarantine policies.[21–25] Much less is known about impacts in lower and middle income countries. Two largescale studies using the government’s electronic immunization records in Pakistan found greater than 50% declines in daily vaccinations during the COVID-19 lockdown.[26,27] A study using administrative records from one hospital in Senegal also documented decreases in vaccine doses given relative to previous year.[25] However, whether immunization coverage recovered after lockdowns were lifted and immunization activities resumed remains unclear.

India recorded its first COVID-19 case in January. As case numbers began to increase, the Indian government announced a nationwide lockdown on March 24, 2020 that barred people from leaving their homes and halted almost all commercial activities and transport services to control virus spread.[28] The nationwide lockdown lasted 10 weeks, until May 31, 2020, after which restrictions continued in areas with high case counts. Although health facilities and critical health services were exempt from the lockdown, immunization outreach sessions were discontinued due to concerns that they may increase the spread of COVID-19.[29] Media reports documented sharp declines in monthly immunizations in April.[30] In late May, the Ministry of Health issued updated guidelines calling for states to resume immunization outreach activities in locations that had prevented or contained COVID-19, with a focus on conducting catch-up immunizations.[31]

Catch-up vaccinations are a critical component of the health system’s response during and after a pandemic, as missed immunizations increase the health risks of unvaccinated children and the population.[32–35] But they make substantial additional demands of already stretched resources.[7,36] They require identification of individuals, groups, or cohorts with missed immunizations, communication to increase awareness and allay fears, additional supply chain management, and intensified catch-up activities, all while continuing routine immunization services and containing pandemic spread.[37] Understanding the extent to which catch-up efforts effectively reached children who had missed their immunizations is critical to adapting and targeting policies and immunization outreach activities in the coming months. However, publicly available administrative data is not sufficient to answer this question. In India, immunization coverage is monitored through the government’s Health Management Information System (HMIS), which collates data from the primary health care system and reports the total number of children immunized each month. Analysis of these data shows that monthly immunizations dropped by 70% nation-wide in April, but then rebounded substantially (supplement table S1, figures S1, S2). In Rajasthan, immunizations in May were 23% higher than pre-lockdown levels. While these aggregate changes provide an important measure of the health system’s recovery, they cannot assess the extent of catch-up among cohorts with missed immunizations or identify groups that remain unvaccinated, for which individual immunization status is required.

This study aimed to use primary survey data to examine disruptions to child immunizations during the COVID-19 lockdown and the extent to which the government’s efforts at catch-up immunization after the lockdown was eased were successful at reaching missed children in low-income households in Rajasthan, India.

## METHODS

### Study Population and Data

We conducted a retrospective observational study using primary data collected through phone surveys to track changes in the immunization status of children that turned one year old between January and October 2020. The study was conducted in Rajasthan, the seventh most populous state in India, and among the states with early confirmed COVID-19 cases.[38] Immunization services, except those provided at health facilities and at birth, were suspended statewide in the first weeks of the lockdown. Active immunization outreach through local health facilities and village camps resumed in May, with particular attention given to ensuring catch-up among missed children, but services remained suspended in areas with high case numbers classified as COVID red zones.[31]

We focused on children in households participating in the Bhamashah Swasthya Bima Yojana (BSBY), a statewide government health insurance program in Rajasthan. BSBY was launched in 2015 and entitled low-income households across the state to free hospital care, including child deliveries, in public and private hospitals. By 2019, over 6 million insurance claims had been filed under the program. We obtained access to administrative data on all insurance claims filed under the program through October 2019. These data included the service provided and the patient’s age, sex, phone number, and residence location. To identify households with at least one child that turned 12 months old between January and October 2020, and therefore eligible for immunizations in the months before and after the COVID-19 lockdown, we identified all claims for a child delivery between January and October 2019. We stratified claims by delivery month and the household’s residence district, and selected random samples of equal size within each stratum to be surveyed by phone using the numbers included in the insurance records. Because we expected a high share of phone numbers in the administrative records to be incorrect or outdated by the time of the survey, we sampled a substantially higher number of households than we expected would be reached.

The survey was conducted between August and early October 2020. Sampled households were randomly assigned to enumerators to avoid enumerator bias. Up to five attempts were made to reach every phone number on record for each household. Households that confirmed having no child born in or after January 2019 and at least 12 months old by the time of the survey were not surveyed. The survey was conducted with an adult that was most knowledgeable about the health of the children in the household and collected immunization histories, as well as data on COVID-related immunization disruptions, contacts with health care workers, demographics, and socioeconomic status. Details of whether or not every immunization on the national schedule was received, as well as the date of their first Measles shot (Measles1) and most recent immunization, were obtained from the child’s immunization card if available. For children whose immunization card was not available at the time of the survey, we asked parents to report immunization details. Because parents have difficulty identifying each immunization and the precise date at which it was delivered, they were asked the total number of injectable immunizations after birth and date and location of the last immunization the child had received.[39]

### Exposure Groups

To analyze changes in immunization status by how affected children were likely to be by the lockdown, we classified them into groups based on their age at the time of the COVID-19 lockdown period. The lockdown took effect in late March and extended through the end of May. The Measles1 injection is the last of the key first-year immunizations and is due at 9-12 months per the national immunization schedule, though historical data shows that most children get it in their ninth month.[40] Therefore, children who turned 12 months before March 2020 should have received all their first-year immunizations before the lockdown and should be least affected. We classified them as “unexposed”. Children who turned 12 months between March and May 2020 (9 months in December – February) may have been affected by lockdown disruptions if they had not already received their first-year immunizations by 11 months and were classified as “partially exposed”. Children who turned 12 months between June and August (9 months in March - May), were due their Measles1 during the worst of the lockdown and were likely to have faced the most severe consequences. They were classified as “heavily exposed”. Finally, children who turned 12 months in September or October (9 months in June or July), after the worst of the lockdown and when the government had resumed immunization activities, were classified as “post-exposure”.

### Outcomes and Measures

The discontinuation of immunization sessions in parts of March and April is expected to have caused children to miss immunizations due that month, but catch-up efforts that began later could have successfully reached these children. To study the extent of missed and catch-up immunizations, we examined the age at which children received their Measles1 immunization. Because parents were unable to reliably remember the name of each immunization over time and the age or date at which it was received, we conducted the analysis of Measles1 for children with an immunization card, which recorded the date for Measles1 if they had received it. Age at Measles1 was calculated as the difference between month of birth and month in which Measles1 was received. We created binary measures for Measles1 immunization received at or before 9 months of age and for Measles1 immunization received at 10 to 12 months. We excluded Measles1 at ages above 12 months to ensure we did not conflate age and exposure effects, as exposed and post-exposure children were younger at the time of survey.

To examine the immunization status of all children by the time of the survey, including those without an immunization card, we created a summary binary measure for whether the child had received all key first-year immunizations, combining data from the immunization card, if available, and parent reports otherwise. Given the difficulties parents face in recalling the names of immunizations, for children without an immunization card we used the total number of immunizations. We excluded BCG, which is due at birth and is typically provided at the delivery facility, and focused on injectable immunizations to avoid confusion between oral immunizations and other medications. Therefore, for children without an immunization card, the completed key first-year immunization status outcome was equal to 1 if the parents reported the child had received at least 4 injectable immunizations and, for children with an immunization card, it was equal to 1 if the Pentavalent1, Pentavalent2, Pentavalent3, and Measles1 immunizations were recorded on the card. Because this measure captured completed first-year immunization status as of the date of the survey, it included the results of catch-up immunization efforts after the lockdown was eased.

### Statistical Methods

To study differences in immunization outcomes across child exposure groups, we ran logistic regressions of each binary immunization outcome on indicators for exposure classification, with “unexposed” children as the reference group. The analysis of timeliness of Measles1 immunization was conducted on the subsample of children with an immunization card. The analysis of completed key first-year immunization status was conducted on all children, combining data from the immunization card and parent reports. Adjusted regressions included binary indicators for survey month, low assets, and low caste as controls. Because differences between children with and without an immunization card may be correlated with immunization status and reporting accuracy, the adjusted regressions of completed key first-year immunization status included an additional binary indicator for whether the child had an immunization card. We also reported the raw probability of having received all key first-year immunizations from the same regressions.

To test whether children across exposure groups were similar in other characteristics, we used pairwise t-tests of child sex, low assets, low parent education, low caste, having an immunization card, and exposure to COVID red zones. To check the sensitivity of the analysis to whether data were obtained from the immunization card or from parent reports, we also estimated the probability of completed key first-year immunization status, and differences by exposure group, separately for immunization status measured from each data source. Since immunizations due at younger ages should be unaffected by the lockdown, we ran similar logistic regressions with receipt of Pentavalent1 and Pentavalent2, which are due at less than 9 months, as the outcomes on indicators for exposure group as a falsification test.

To examine whether lockdown exposure had differential effects on immunization status across subgroups, we estimated the change in probability of completed key first-year immunization status for heavily exposed relative to unexposed children for each subgroup. These probabilities were estimated after running logistic regressions of the completed immunization on the interaction of exposure group and each subgroup characteristic with controls for survey month and having an immunization card. Binary subgroups were defined for female sex, low assets (below median index score), at or below 12 years of parent schooling, low caste, and residence in a COVID red zone. The asset index was computed as the standardized first component of a principal component analysis of a list of assets. Households belonging to a scheduled caste or tribe group were classified as low caste. All analysis was conducted using Stata version 16.[41]

## RESULTS

### Survey Sample

Of the 5,102 households sampled for survey, 3,418 (67%) were reached by phone and 2,248 households (44.1%) confirmed having a child born in or after January 2019 and at least 12 months old by the time of the survey in the household (supplement table S2). The primary reasons for being unable to reach the household were that the phone number was invalid, outside coverage, or switched off, which was most likely due to households changing their phone numbers over time or possibly to numbers being entered incorrectly in the insurance records. 92.6% of eligible households consented to participating in the study, resulting in a final study sample of 2,081 households and 2,144 children aged at least 12 months old. Surveyed households are similar in pre-survey characteristics to those not surveyed (supplement table S3).

Table 1 presents descriptive statistics for children included in the study. 52% of children were male and 26% were in lower caste households. 1,287 (60%) children had an immunization card available. Children with an immunization card available were less likely to be male and poor (supplement table S4). Recent contacts with the public primary health care system were substantial: 82% of children had seen a health worker since the imposition of the lockdown (table 1). For 31% of children, parents reported experiencing a COVID-related disruption to their immunizations. Fear of contracting COVID-19 was the most common reported cause of disruptions (20%), followed by canceled immunization sessions (15%), and travel barriers that made it difficult to reach the session (10%). 19% of children resided in districts classified as COVID red zones throughout April and May. Overall, 21% of children in the study sample were classified as unexposed to the lockdown, 34% and 37% were classified as partially exposed and heavily exposed, and 9% turned 12 months in September or later and were classified as post-exposure. Children across exposure groups were similar in terms of sex, assets, caste, parental education, and COVID-zone classification (supplement table S5). By construction, unexposed children were oldest (median 19 months) and post-exposure children were youngest (median 12 months) by the time of the survey.

**Table 1:** Characteristics of Study Sample. The study included 2,144 children. Children were classified as low caste if their household belonged to a scheduled caste or tribe and low parent education if neither parent had completed more than 12 years of schooling. Healthcare worker includes Accredited Social Health Activist, Anganwadi Worker or Auxiliary Nurse Midwife, the three frontline primary health workers. COVID red zones are districts classified by the government as having high COVID-19 cases in late April and May.

|  | Number | Percent |
| --- | --- | --- |
| Male | 1,122 | 52 |
| Low caste | 556 | 26 |
| Low parent education | 1,157 | 54 |
| Has immunization card | 1,287 | 60 |
| Seen health worker since lockdown | 1,652 | 82 |
| Any COVID-related immunization disruption | 649 | 31 |
| Avoided due to COVID fear | 407 | 20 |
| Immunization camp not held | 315 | 15 |
| Travel barriers | 215 | 10 |
| Residence in COVID red zone | 414 | 19 |
| **Exposure Groups** |  |  |
| Unexposed (12mo before Mar) | 443 | 21 |
| Partially exposed (12mo Mar-May) | 722 | 34 |
| Heavily exposed (12mo Jun-Aug) | 796 | 37 |
| Post-exposure (12mo Sep-Oct) | 183 | 9 |
| Observations | 2,144 |  |

### Timeliness of Measles1 Immunizations

Figure 1 presents adjusted odds ratios for receiving Measles1 immunization at or before 9 months and between 10-12 months by exposure group. Unadjusted estimates are in supplement table S6. Heavily exposed children experienced a shift in the timing of their Measles1 immunization compared to both unexposed and post-exposure children. They were significantly less likely to have been immunized at or before 9 months than unexposed children (OR 0.550, p=0.004, 95% CI 0.367 to 0.824) and were significantly more likely to get their immunization later, between 10-12 months (OR 1.761, p=0.004, 95% CI 1.196 to 2.591), suggesting that the efforts to conduct catch-up immunizations after the lockdown reached many of these children at later ages. Post-exposure children showed no differences in timeliness of Measles1 immunizations relative to unexposed children, indicating that children turning 9 months in June or later returned to pre-lockdown immunization schedules. The complete breakdown by age in supplement figure S3 shows that 51% of unexposed children had already received their Measles1 at or before 9 months, this dropped to 33% among heavily exposed children, and increased to 45% among post-exposure children. However, 33% of heavily exposed children were immunized at 11 or 12 months, compared to 14% of unexposed and 19% of post-exposure children, indicating that a substantial share of heavily exposed children was immunized between May and July, when immunization activities resumed.

**Figure 1:**
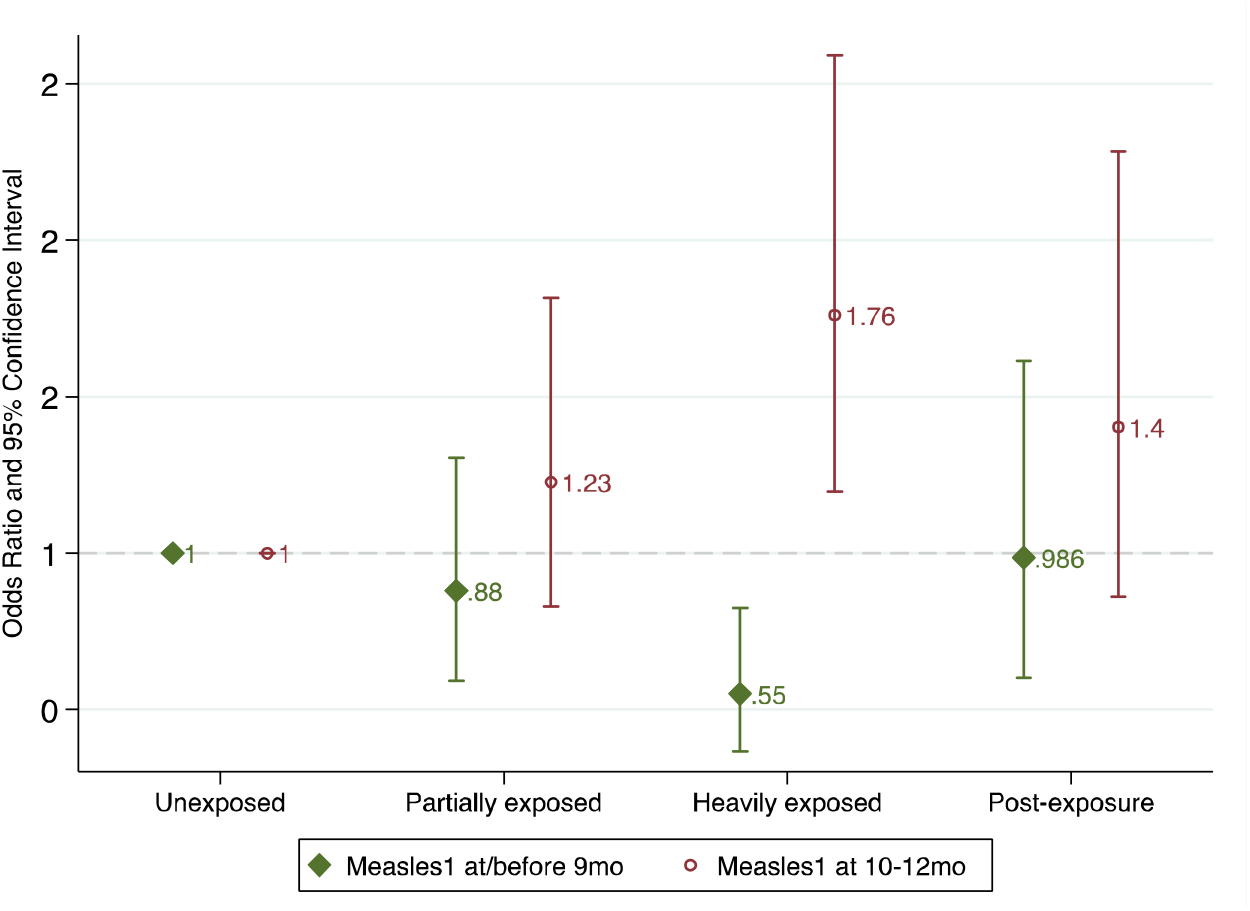
Measles1 Immunization at or before 9 months and between 10-12 months by Lockdown Exposure. The figure presents odds ratios with 95% confidence intervals from adjusted logistic regressions of Measles1 received at or before 9 months and received between 10 and 12 months on indicators for exposure group, with unexposed children that turned 12 months old before March 2020 as the reference group. The sample was restricted to children with an immunization card and date of Measles1 recorded, if received. Indicators for survey month, low assets, and low caste were included as controls. Details and unadjusted estimates are in supplemental table S6.

### Completed First-Year Immunization Status

Figure 2 presents the unadjusted probability of having completed all key first-year immunizations by the time of the survey across exposure groups, combining children with and without an immunization card. Among unexposed children, who turned 12 months old in February 2020 or earlier, 74.5% (95% CI 70.4%-78.6%) had received all their key first-year immunizations. Coverage dropped to 70.4% (95% CI 67.0% to 73.7%) among partially exposed children and 64.1% (95% CI 60.7% to 67.4%) among heavily exposed children, a 10.4pp or 14% decrease relative to unexposed children. However, among post-exposure children coverage increased to 71.0% (95% CI 64.5% to 77.6%), slightly lower than the level for the unexposed and 6.9pp higher than the heavily exposed. The complete breakdown by birth cohort shows a fairly steady decline in coverage across children classified as partially and heavily exposed, followed by a rebound amongst the post-exposed (supplement figure S4, table S7).

**Figure 2:**
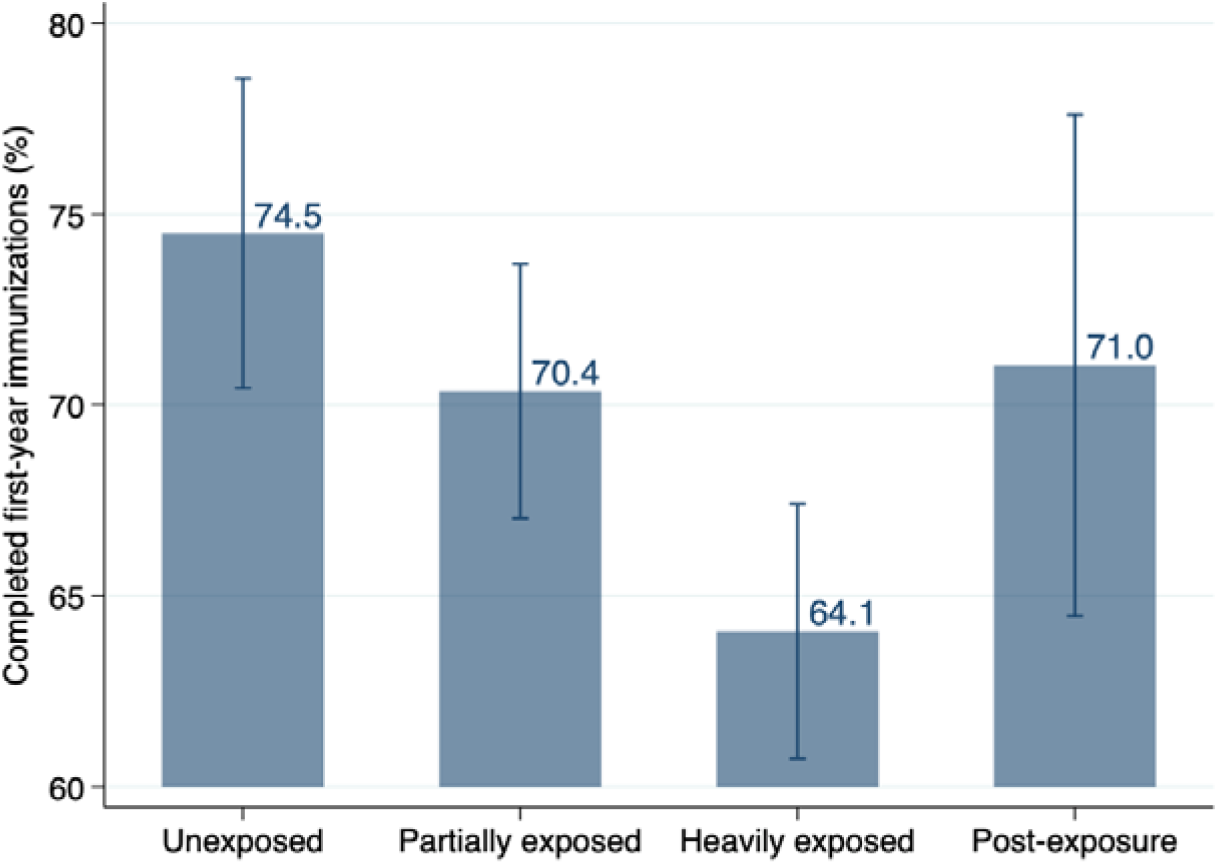
Probability of Completed First-Year Immunizations by Lockdown Exposure. The figure presents unadjusted probabilities of having received all key first-year immunizations by lockdown exposure group with 95% confidence intervals. The outcome is equal to one if the child received Pentavalent1, Pentavalent2, Pentavalent3, and Measles1 on the immunization card or at least 4 injectable immunizations per parent reports, if the immunization card was unavailable. The complete breakdown by age cohort is in supplemental figure S4.

Table 2 presents regression results directly comparing completed first-year immunization across exposure groups relative to unexposed children. The results confirm the patterns in Figure 2. Heavily exposed children were significantly less likely to have received all their key first-year immunizations by the time of the survey than unexposed children (adjusted OR 0.624, p=0.001, 95% CI 0.478 to 0.816), while post-exposure children rebounded, with an odds ratio that was higher than heavily exposed children and not significantly different from those of unexposed children (adjusted OR 0.825, p=0.354, 95% CI 0.549 to 1.240). Part of the decline in coverage between unexposed and heavily exposed children may be due to the lower age of the latter by the time of the survey. However, comparison with the higher estimate for post-exposure children, who were youngest at the time of the survey, relative to heavily exposed children indicates that at least half of this decline cannot be explained by age.

**Table 2:** Difference in Completed First-Year Immunization Status by Lockdown Exposure. The table presents odds ratios from logistic regressions of a binary indicator for having received all key first-year immunizations on indicators for exposure group, with unexposed children that turned 12 months old before March 2020 as the reference group. The outcome is equal to one if the child received Pentavalent1, Pentavalent2, Pentavalent3, and Measles1 on the immunization card or at least 4 injectable immunizations per parent reports, if the immunization card was unavailable. Adjusted regressions include indicators for survey month, low assets, low caste, and whether the child had an immunization card as controls.

|  | Unadjusted |  |  | Adjusted |  |  |
| --- | --- | --- | --- | --- | --- | --- |
|  | OR | p-value | 95% Confidence Intervals | OR | p-value | 95% Confidence Intervals |
| Unexposed | Ref |  |  | Ref |  |  |
| Partially exposed | 0.813 | 0.128 | (0.623 to 1.061) | 0.804 | 0.122 | (0.610 to 1.060) |
| Heavily exposed | 0.611 | 0.000 | (0.472 to 0.790) | 0.624 | 0.001 | (0.478 to 0.816) |
| Post-exposure | 0.840 | 0.374 | (0.572 to 1.233) | 0.825 | 0.354 | (0.549 to 1.240) |
| Observations | 2,144 |  |  | 2,144 |  |  |

In supplement table S8 we examined the sensitivity of the results to whether data were obtained from the immunization card or parent reports. The probability of completed first year immunization was higher among children with an immunization card in every exposure group, which may reflect underreporting by parents or higher immunization coverage among children with a card. However, differences in immunization coverage across exposure groups showed similar patterns across the two data sources: large decreases among heavily exposed children relative to unexposed and a rebound among post-exposure children (supplement table S8). Falsification tests examining immunizations due prior to the ninth month, that should be unaffected by exposure to the lockdown, did not show any differences by exposure group (supplement table S9).

### Subgroup Analysis

Among unexposed children, the probability of completed first year immunization status was lower among those in less educated (70.1% vs 79.0%), poorer (71.5% vs 77.1%), and lower caste (72.6% vs 74.8%) households (figure 3). Comparing differences between heavily exposed and unexposed children, declines were also larger for each of these subgroups than for their more educated, wealthier, and higher caste counterparts, although the subgroup comparisons were not significant at the 95% level (figure 3, supplement table S10). The largest difference was by education: the decline among heavily exposed children was 5.8pp (p=0.101) in more educated households and 13.0pp (p=0.000) in less educated households (7.2pp difference in declines, p=0.160). Children residing in COVID red zones also experienced a larger decline (17.9pp, p=0.002) than those in other areas (7.7pp, p=0.009; 10.2pp difference; p=0.109). For most groups, coverage among post-exposure children was similar to that among unexposed children, but levels remained lower in COVID red zones (supplemental table S10).

**Figure 3:**
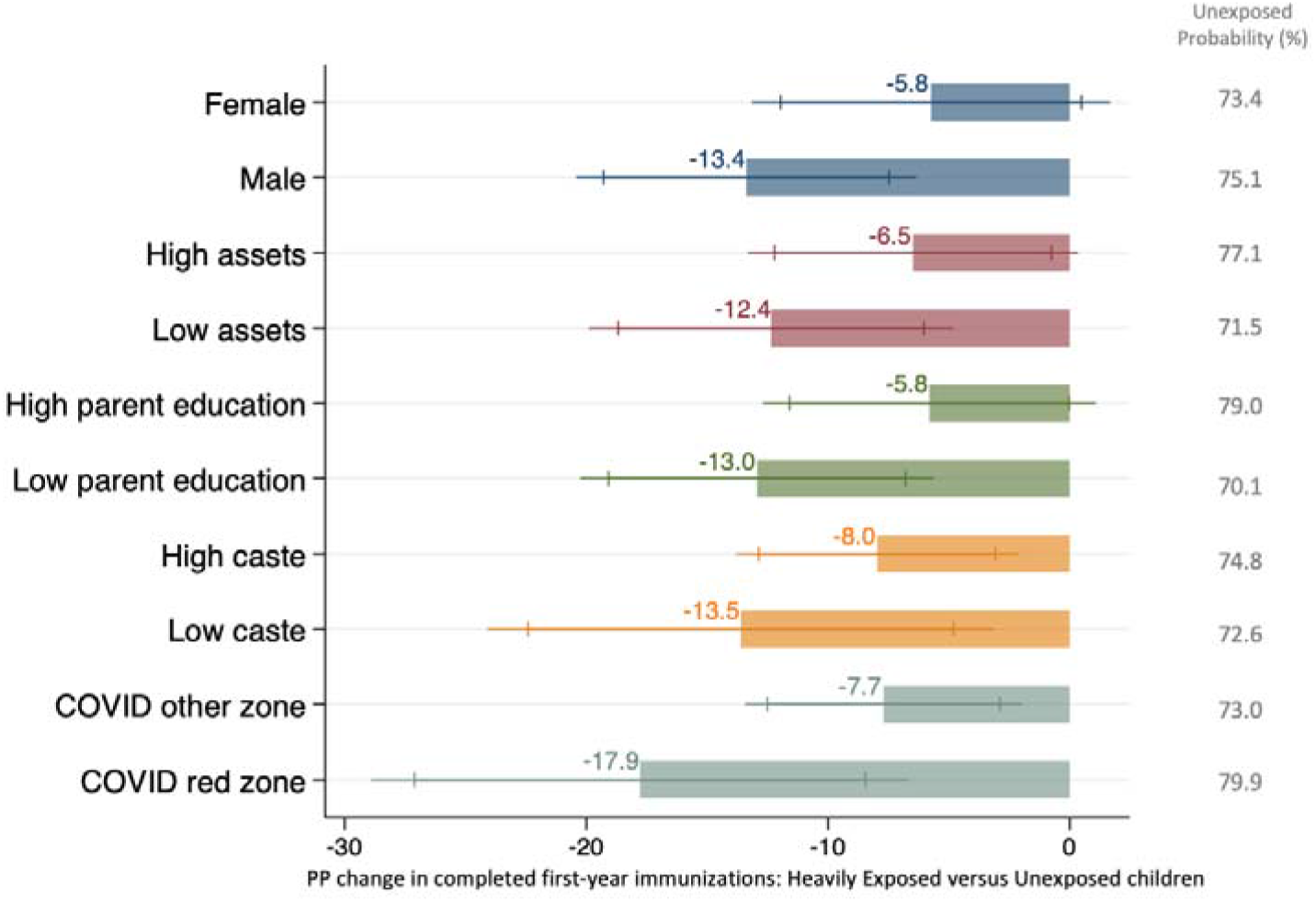
Declines in Probability of Completed First-Year Immunization by Subgroup. The figure presents the percentage point change in probability of having received all key first-year immunizations between heavily exposed and unexposed children, by subgroup. The column on the right indicates the probability of completed first-year immunization status among unexposed children for each subgroup. Estimates were adjusted for survey month and whether the child had an immunization card. Details are in supplemental table S10.

## DISCUSSION

Using primary data collected through phone surveys on the immunization status of children that turned one year old between January and October 2020, this study found substantial changes in the timeliness of child immunizations. Children due the Measles1 immunization between March and May, during the strictest lockdown, were significantly less likely to receive it at or before 9 months than older cohorts, and more likely to have received the Measles1 immunization at older ages, providing evidence that immunization outreach after the lockdown was eased was successful at reaching many of them.

Nevertheless, catch-up efforts did not fully offset the lapses in immunization. We measured the likelihood of having completed all key first-year immunizations at the time of the survey, 4-5 months after the lockdown was eased, across all exposure groups. Children due immunizations during the lockdown had a significantly lower likelihood of completed first-year immunizations at the time of the survey, when they were about 14 months old, than unexposed children. In contrast, among children that were due their immunizations in June or later, after the lockdown eased, completed immunization was not significantly different from pre-lockdown levels. This finding of a rebound among the youngest cohort in our study is important for several reasons. First, it suggests immunization activities may already be on an upward trajectory and returning to pre-lockdown schedules, although this progress will need continuous monitoring. Second, while the lower immunization coverage among heavily exposed relative to unexposed children may reflect the younger age of the former, comparing them with the younger post-exposure children clarifies that age cannot explain the decline. Instead, it provides an alarming sign that, despite the resumption of immunization services, children most exposed to the lockdown were not fully reached through catch-up efforts, even as immunizations among younger cohorts increased, and may be at risk of being missed without careful tracking and targeted interventions.

Subgroup analysis found that children in less educated, poorer, and lower caste households had lower pre-COVID levels of immunization, consistent with prior evidence on the socioeconomic predictors of delayed and incomplete vaccination in India.[42,43] Worryingly, these subgroups also experienced somewhat larger declines during the lockdown that catch-up efforts had not addressed by the time of this study. These disparities may reflect lower trust or awareness of the importance of immunization among households, but also the inadequate reach of the health system among these populations. An effective post-pandemic response will need to ensure these groups are not left out. We also found larger declines in immunization coverage among children in COVID red zones that are unsurprising, as services in these areas were discontinued through May and may still be suspended.[29] However, the risk of vaccine preventable infectious disease outbreaks in these areas needs to be weighed against the risk of COVID-19 spread through immunization outreach.[20]

Our findings provide a snapshot of the immunization situation 4-5 months after India’s nationwide lockdown was eased. Immunization status may continue to improve if catch-up efforts to reach children with pending immunizations are successful at addressing both demand and supply side constraints. Nevertheless, our findings offer an early understanding of how immunization status has evolved after the recommencement of services, and the success and shortfalls in coverage that policy efforts must focus on. It is also important to note that, per the HMIS, post-lockdown increases in immunizations were larger in Rajasthan than in other states, suggesting that catch-up may have been more successful in Rajasthan and the share of children with incomplete vaccinations may be much higher elsewhere in India (supplement table S1).

An important implication of this study is that successful catch-up requires careful monitoring of the immunization status of individuals and groups.[36,37] Monitoring metrics that track total numbers of children immunized each month cannot identify the specific groups that have fallen behind that catch-up efforts must target. In supplementary analysis of the government’s administrative HMIS data for Rajasthan, we found monthly full immunizations declined from 113,190 before the lockdown to 14,514 in April (−87%) but then increased to 155,416 (+37%) and 141,389 (+25%) in May and June (supplement table S1). Based on the HMIS data, the Rajasthan government announced that it had exceeded its immunization targets in May and June.[44] However, aggregate trends may increase due to seasonal fluctuations in immunization demand rather than successful catch-up. We found that correcting for historical trends reduced these estimates to 23% in May and -4% in June. Furthermore, it is impossible to ascertain whether these aggregate increases reflect expanded immunization coverage among younger cohorts or successful catch-up among older cohorts already in the system, nor whether specific subgroups were left out. Domestic migration, which was substantial after the lockdown in India, further complicates interpretation of absolute numbers.[45] Our study shows that, while there were indeed increases in post-lockdown immunization outreach and coverage, they did not fully offset immunizations missed during the lockdown, particularly among groups that are typically underserved by the health system.

While strengthening the supply of services is critical, catch-up efforts also need to address the demand side. Consistent with evidence no COVID-19 and prior outbreaks, we found that fear of contracting COVID was a commonly reported reason for missed immunizations.[6,8,9,12] Public health policies will need to allay these fears by building trust in public primary care services, clearly communicating the importance of continuing immunizations even during a pandemic, and ensuring that attending immunization camps does not increase the risk of COVID infection.

This study has several limitations. First, the sample is households with an institutional child delivery under the BSBY government health insurance program and findings may not be representative of Rajasthan’s population. Given that BSBY targets low-income households across the state, this population is of particular policy interest, but it may exclude the poorest households that underutilize health facilities. Second, although phone surveys allowed us to rapidly survey a large sample of households at a time when in-person data collection was impossible, because phone numbers were obtained from administrative records from a year ago, this resulted in substantial attrition in the survey. We expected this at the start of the study and oversampled households for survey. Surveyed households were not significantly different from those we were unable to reach on characteristics available in the administrative records, increasing confidence that attrition did not bias our study sample. Third, the study relied on data from the immunization card and parent reports of immunization status, both of which may be subject to error. We found that estimates of levels of completed immunization coverage from immunization cards were typically higher than those from parent reports, which could reflect biases in either data source, but also real differences in immunization status across these groups. Largescale validations of recall in low income contexts have found that parent-reported immunization status is reliable and including it can yield more representative estimates of coverage than health cards alone.[46] Furthermore, our analysis of differences in immunization status across exposure groups showed similar patterns across children with and without an immunization card, suggesting any potential biases were not likely to be differential across exposure groups.

Despite substantial gains in immunization coverage over the last decade, India accounts for 37% of global measles deaths and Rajasthan has lower immunization coverage than the rest of the country.[47–49] The state resumed immunization activities quickly and effectively after the lockdown and reversed much of the effect of the pandemic on vaccination status. However, without careful tracking to identify the specific individuals and groups that missed immunizations, and targeted efforts prioritizing them for catch-up, a large share of children may remain unvaccinated, turning back decades of progress against infectious diseases.

## Data Availability

Data are available from the authors upon request.

## Acknowledgements

We would like to thank Putul Gupta for management support on this project and the team of surveyors that helped collect the data.

## Contributors

RJ and PD developed the research question and study design. MP and AC oversaw data collection and did the data cleaning. MP, CF, and RJ conducted the statistical analysis. RJ wrote the initial manuscript draft. All authors revised drafts and approved of its final version.

## Ethics

This study received ethical clearance from the institutional review boards at the Institute for Financial Management and Research (IFMR) in India and Stanford University in the US (protocol no. 41683). Participants gave oral consent before taking part in the survey.

