## Supplemental Materials for "COVID-19 RELATED IMMUNIZATION DISRUPTIONS IN RAJASTHAN, INDIA: A RETROSPECTIVE OBSERVATIONAL STUDY"

#### A. HMIS Analysis

To supplement our survey results with data from across Rajasthan and all of India, we used publicly available immunization data from the government's Health Management Information System (HMIS), which collects monthly reports from the primary health system across the state. The collated data are publicly available online.[1,2] These data are based on paper records kept by frontline health workers and may have quality issues.[3] Nevertheless, they provide the most comprehensive measures of immunization activities in India and are used by policymakers for decision-making.

The outcomes available in the HMIS that we studied were immunization sessions, children that received each of the key first-year immunizations (BCG, Pentavalent1, Pentavalent2, Pentavalent3, Measles1), and children 9-11 months that were fully immunized. Figure S1 presents the monthly fully immunized children in Rajasthan from November 2019 to June 2020 (the last month of complete data), and for the same months in the previous 2 years. Numbers are relatively stable between November and February each year; but there is also evidence of seasonal trends. Therefore, to calculate changes over the lockdown period, we used data from November 2019 to June 2020 and November 2018 to June 2020 for the analysis. We pooled November to February as the reference and reported monthly percent changes relative to it. To account for historical trends, we also reported monthly changes in 2020 after adjusting for the change over the same period in the previous year: e.g. Change March2020 = 1 – (March2020/Reference2020)/(March2019/Reference2019). We also calculated changes separately for districts that were classified as COVID red zones between late April and mid-May and all other districts, because immunization outreach was suspended in the former. To examine Rajasthan's experience relative to the rest of the country, we conducted the same exercise for fully immunized children aged 9-11 months for all of India, as well as Bihar, Madhya Pradesh, and Uttar Pradesh.

Complete data are presented in supplement table S1 and historically adjusted changes in fully immunized children are presented in supplement figure S2. Monthly full immunizations in Rajasthan declined from 113,190 in the reference period to in 84,037 in March and 14,514 in April (-26% and -87% change), but numbers in May increased to 155,416 (37%). However, adjusting for historical trends increased the estimated decline to -45% in March and reduced the reversal in May to 23%. The remaining discussion focuses on adjusted estimates. Trends were similar across all key first year immunizations, except for BCG, which experienced a smaller decline of 41% in April, possibly due to the government's policy to continue immunizations provided during births at health facility through the lockdown. The rebound in May was substantially smaller in districts that had been classified as COVID red zones. Supply disruptions were a major explanatory factor, as monthly immunization sessions also decreased substantially in April. The recovery in May (23%) and June (-4%) was larger in Rajasthan than for India (-23% and -31%), suggesting that Rajasthan was among the more successful states in resuming immunization activities soon after the lockdown was eased.

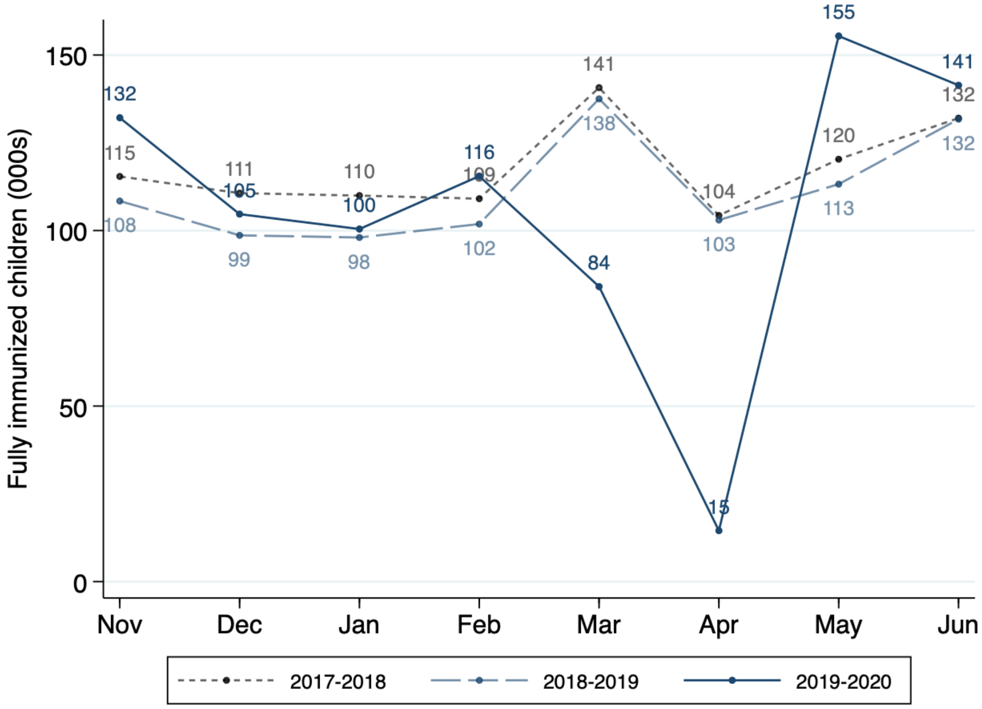

**Figure S1: Monthly HMIS Trends of Fully Immunized Children Aged 9-11 Months in Rajasthan**
The figure presents the monthly number of fully immunized children aged 9-11 months between November and June for the current and 2 previous years in Rajasthan as reported in the HMIS data.

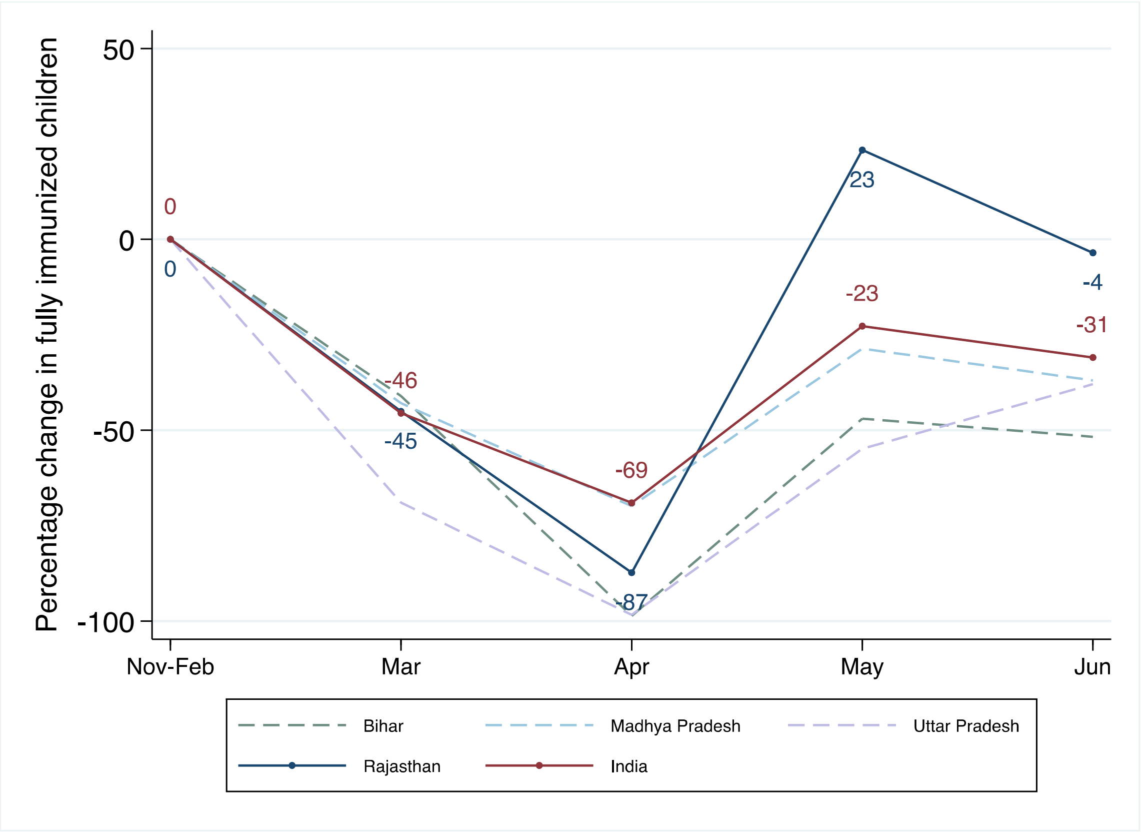

**Figure S2: HMIS Changes in Monthly Fully Immunized Children Aged 9-11 Months in 2020**
The graph presents monthly changes in fully immunized children 9-11 months between March and June relative to the November 2019 to February 2020 reference period, adjusting for changes over the same period in the previous year. The blue solid line is changes in Rajasthan. For comparison, the graph presents changes in all of India (red solid line), as well as Bihar, Madhya Pradesh and Uttar Pradesh (dashed lines). Immunizations in Rajasthan dropped by 45% in March relative to pre-lockdown levels and by 87% in April, but increased by 23% in May. Both the decline and rebound were larger than in the all of India.

| PANEL A: NOVEMBER 2019 TO JUNE 2020 | | | | | | | | | | | | | | | | |
|---|---|---|---|---|---|---|---|---|---|---|---|---|---|---|---|---|
| | **Nov-Feb Mean** | **March** | | | **April** | | | **May** | | | **June** | | | **March-June Mean** | | |
| | Number | Number | % Ref | % Adj | Number | % Ref | % Adj | Number | % Ref | % Adj | Number | % Ref | % Adj | Number | % Ref | % Adj |
| **RAJASTHAN** | | | | | | | | | | | | | | | | |
| Fully Immunized 9-11mo | 113,190 | 84,037 | -26 | -45 | 14,514 | -87 | -87 | 155,416 | 37 | 23 | 141,389 | 25 | -4 | 98,839 | -13 | -28 |
| Immunization sessions | 65,570 | 51,934 | -21 | -20 | 7,433 | -89 | -89 | 55,884 | -15 | -15 | 62,331 | -5 | -5 | 44,396 | -32 | -32 |
| BCG | 119,352 | 98,716 | -17 | -21 | 55,249 | -54 | -41 | 104,257 | -13 | 1 | 103,417 | -13 | -8 | 90,410 | -24 | -17 |
| Pentavalent1 | 127,436 | 88,930 | -30 | -26 | 16,167 | -87 | -84 | 140,996 | 11 | 43 | 113,771 | -11 | 11 | 89,966 | -29 | -14 |
| Pentavalent2 | 125,604 | 95,264 | -24 | -27 | 13,394 | -89 | -87 | 97,614 | -22 | -5 | 133,141 | 6 | 33 | 84,853 | -32 | -21 |
| Pentavalent3 | 123,632 | 95,864 | -22 | -30 | 13,159 | -89 | -88 | 101,468 | -18 | -5 | 103,983 | -16 | -2 | 78,618 | -36 | -31 |
| Measles1 | 119,214 | 86,600 | -27 | -46 | 15,251 | -87 | -87 | 161,833 | 36 | 22 | 147,304 | 24 | -4 | 102,747 | -14 | -29 |
| **COVID Red Zone** | | | | | | | | | | | | | | | | |
| Fully Immunized 9-11mo | 28,795 | 20,236 | -30 | -44 | 2,315 | -92 | -92 | 34,062 | 18 | -0 | 34,411 | 20 | -12 | 22,756 | -21 | -37 |
| Immunization sessions | 16,438 | 13,388 | -19 | -18 | 1,620 | -90 | -90 | 12,411 | -25 | -26 | 14,399 | -12 | -12 | 10,454 | -36 | -37 |
| **COVID Other Zone** | | | | | | | | | | | | | | | | |
| Fully Immunized 9-11mo | 84,395 | 63,801 | -24 | -45 | 12,199 | -86 | -86 | 121,354 | 44 | 32 | 106,978 | 27 | -0 | 76,083 | -10 | -25 |
| Immunization sessions | 49,132 | 38,546 | -22 | -20 | 5,813 | -88 | -88 | 43,473 | -12 | -11 | 47,932 | -2 | -2 | 33,941 | -31 | -30 |
| **INDIA** | | | | | | | | | | | | | | | | |
| Fully Immunized 9-11mo | 2,059,236 | 1,805,660 | -12 | -46 | 664,406 | -68 | -69 | 1,880,668 | -9 | -23 | 1,822,266 | -12 | -31 | 1,543,250 | -25 | -42 |
| Immunization sessions | 1,081,912 | 981,357 | -9 | -14 | 396,937 | -63 | -62 | 835,248 | -23 | -27 | 842,482 | -22 | -26 | 764,006 | -29 | -32 |

| PANEL B: NOVEMBER 2018 TO JUNE 2019 | | | | | | | | | | | |
|---|---|---|---|---|---|---|---|---|---|---|---|
| | **Nov-Feb Mean** | **March** | | **April** | | **May** | | **June** | | **March-June Mean** | |
| | Number | Number | % Ref | Number | % Ref | Number | % Ref | Number | % Ref | Number | % Ref |
| **RAJASTHAN** | | | | | | | | | | | |
| Fully Immunized 9-11mo | 101,733 | 137,524 | 35 | 103,000 | 1 | 113,214 | 11 | 131,732 | 29 | 121,368 | 19 |
| Immunization sessions | 65,208 | 64,258 | -1 | 65,270 | 0 | 65,138 | -0 | 65,082 | -0 | 64,937 | -0 |
| BCG | 118,575 | 124,692 | 5 | 93,642 | -21 | 102,086 | -14 | 112,112 | -5 | 108,133 | -9 |
| Pentavalent1 | 128,418 | 120,833 | -6 | 103,202 | -20 | 99,142 | -23 | 103,019 | -20 | 106,549 | -17 |
| Pentavalent2 | 125,591 | 130,483 | 4 | 104,631 | -17 | 102,496 | -18 | 99,761 | -21 | 109,343 | -13 |
| Pentavalent3 | 121,002 | 134,112 | 11 | 110,575 | -9 | 104,963 | -13 | 103,874 | -14 | 113,381 | -6 |
| Measles1 | 108,569 | 146,556 | 35 | 109,168 | 1 | 120,335 | 11 | 139,581 | 29 | 128,910 | 19 |
| **COVID Red Zone** | | | | | | | | | | | |
| Fully Immunized 9-11mo | 25,607 | 32,159 | 26 | 25,386 | -1 | 30,385 | 19 | 34,936 | 36 | 30,716 | 20 |
| Immunization sessions | 15,863 | 15,843 | -0 | 15,841 | -0 | 16,144 | 2 | 15,875 | 0 | 15,926 | 0 |
| **COVID Other Zone** | | | | | | | | | | | |
| Fully Immunized 9-11mo | 76,126 | 105,365 | 38 | 77,614 | 2 | 82,829 | 9 | 96,796 | 27 | 90,651 | 19 |
| Immunization sessions | 49,346 | 48,415 | -2 | 49,429 | 0 | 48,994 | -1 | 49,207 | -0 | 49,011 | -1 |
| **INDIA** | | | | | | | | | | | |
| Fully Immunized 9-11mo | 1,657,961 | 2,670,398 | 61 | 1,729,129 | 4 | 1,959,982 | 18 | 2,125,477 | 28 | 2,121,246 | 28 |
| Immunization sessions | 985,967 | 1,045,832 | 6 | 953,180 | -3 | 1,045,400 | 6 | 1,037,008 | 5 | 1,020,355 | 3 |

**Table S1: HMIS Monthly Immunization Trends in Rajasthan and India**

The table presents immunization trends from the government's HMIS data for November 2019 to June 2020 in Panel A and November 2018 to June 2019 in Panel B for comparison. The monthly average in the November to February period is the reference period in each year. The table present monthly numbers, the percentage change relative to the reference mean for the same year (% Ref), and the historically adjusted percentage change in 2020 that accounts for the change over the same period in the previous year (% Adj).

### B. Survey Analysis

| | Percent | Count | Observations |
|---|---|---|---|
| Sampled households | | 5,102 | 5,102 |
| Reached by phone | 0.670 | 3,418 | 5,102 |
| Reached and eligible children confirmed | 0.441 | 2,248 | 5,102 |
| Consented to survey | 0.926 | 2,081 | 2,248 |
| Surveyed households | | | 2,081 |
| Children in Study | | | 2,144 |

**Table S2: Survey Status**

This table presents complete statistics on the full sample of households sampled. The primary reasons for being unable to reach a household were that the phone number was invalid, outside coverage, or switched off, which was most likely due to households changing their phone numbers over time or possibly to numbers being entered incorrectly in the insurance records. Reached households which confirmed having at least one child aged 12-month-old or more living in the household were eligible for the survey. Children in study are the children aged at least 12 months old for whom we collected immunization data.

| | (1) Not Surveyed | (2) Surveyed | (3) Overall | (4) (1) vs. (2), p-value |
|---|---|---|---|---|
| Age of mother at delivery | 25.86 | 25.62 | 25.76 | 0.11 |
| | (0.10) | (0.11) | (0.07) | |
| Residence village population | 3974.52 | 4157.70 | 4048.92 | 0.76 |
| | (362.99) | (505.71) | (297.71) | |
| Residence village low-caste share | 0.37 | 0.34 | 0.36 | 0.00 |
| | (0.01) | (0.01) | (0.00) | |
| Village has a public health facility | 0.69 | 0.67 | 0.68 | 0.21 |
| | (0.01) | (0.01) | (0.01) | |
| Village distance to nearest town | 26.25 | 26.85 | 26.50 | 0.39 |
| | (0.44) | (0.54) | (0.34) | |
| Village distance to disctrict HQ | 59.36 | 60.13 | 59.68 | 0.51 |
| | (0.74) | (0.87) | (0.56) | |
| COVID red zone | 0.20 | 0.19 | 0.20 | 0.73 |
| | (0.01) | (0.01) | (0.01) | |
| Observations | 3,021 | 2,081 | 5,102 | |

**Table S3: Household Characteristics by Survey Status**
The table compares pre-survey characteristics of households that were sampled but not surveyed and those successfully surveyed. Columns 1-3 report group means and standard errors in parentheses. Column 4 reports the p-values from t-tests of the difference in means of each variable across surveyed and not surveyed households. We obtained age of mother at delivery from the insurance administrative claims data. Village characteristics were obtained from the 2011 Population Census, which we were able to merge with our survey data using household residence village.

| | (1) Parents Report | (2) Immunization Card | (3) Overall | (4) (1) vs. (2), p-value |
|---|---|---|---|---|
| Male | 0.55 | 0.51 | 0.52 | 0.09 |
| | (0.02) | (0.01) | (0.01) | |
| Low caste | 0.24 | 0.27 | 0.26 | 0.07 |
| | (0.01) | (0.01) | (0.01) | |
| Low asset | 0.55 | 0.47 | 0.50 | 0.00 |
| | (0.02) | (0.01) | (0.01) | |
| Low parent education | 0.55 | 0.53 | 0.54 | 0.40 |
| | (0.02) | (0.01) | (0.01) | |
| COVID red zone | 0.19 | 0.19 | 0.19 | 0.95 |
| | (0.01) | (0.01) | (0.01) | |
| Observations | 857 | 1287 | 2144 | |
| Proportion | 0.40 | 0.60 | 1.00 | |

**Table S4: Characteristics of Children With and Without an Immunization Card**
The table compares characteristics of children with an immunization card and those without one, where full immunization status was based on parent reports. Columns 1-3 report group means and standard errors in parentheses. Column 4 reports the p-values from pairwise t-tests of the differences of the means of each variable across the two sub-groups.

| | (1) Unexposed | (2) Partially exposed | (3) Heavily exposed | (4) Post-exposure | (5) (1) vs. (2), p-value | (6) (1) vs. (3), p-value | (7) (1) vs. (4), p-value | (8) (2) vs. (3), p-value | (9) (2) vs. (4), p-value | (10) (3) vs. (4), p-value |
|---|---|---|---|---|---|---|---|---|---|---|
| Male | 0.54 | 0.53 | 0.50 | 0.55 | 0.64 | 0.20 | 0.91 | 0.35 | 0.65 | 0.30 |
| | (0.02) | (0.02) | (0.02) | (0.04) | | | | | | |
| Has immunization card | 0.61 | 0.62 | 0.58 | 0.61 | 0.82 | 0.28 | 0.95 | 0.13 | 0.91 | 0.40 |
| | (0.02) | (0.02) | (0.02) | (0.04) | | | | | | |
| Low asset | 0.48 | 0.49 | 0.51 | 0.51 | 0.60 | 0.23 | 0.42 | 0.45 | 0.64 | 1.00 |
| | (0.02) | (0.02) | (0.02) | (0.04) | | | | | | |
| Low caste | 0.23 | 0.26 | 0.28 | 0.24 | 0.15 | 0.04 | 0.69 | 0.49 | 0.53 | 0.29 |
| | (0.02) | (0.02) | (0.02) | (0.03) | | | | | | |
| Low parent education | 0.53 | 0.55 | 0.53 | 0.57 | 0.52 | 0.96 | 0.39 | 0.41 | 0.65 | 0.34 |
| | (0.02) | (0.02) | (0.02) | (0.04) | | | | | | |
| COVID red zone | 0.20 | 0.19 | 0.20 | 0.19 | 0.62 | 0.91 | 0.83 | 0.66 | 0.89 | 0.88 |
| | (0.02) | (0.01) | (0.01) | (0.03) | | | | | | |
| Observations | 443 | 722 | 796 | 183 | | | | | | |
| Proportion | 0.21 | 0.34 | 0.37 | 0.09 | | | | | | |

**Table S5: Child Characteristics Across Exposure Groups**
The table compares characteristics of children classified into the four exposure groups. Columns 1-4 report group means and standard errors in parentheses. Columns 5-10 report the p-values from pairwise t-tests of the differences of the means of each variable across exposure groups.

| | Unadjusted | | | | | | Adjusted | | | | | |
|---|---|---|---|---|---|---|---|---|---|---|---|---|
| | Measles1 at/before 9mo | | | Measles1 10-12mo | | | Measles1 at/before 9mo | | | Measles1 10-12mo | | |
| | OR | p-value | 95% Confidence Interval | OR | p-value | 95% Confidence Interval | OR | p-value | 95% Confidence Interval | OR | p-value | 95% Confidence Interval |
| Unexposed | Ref | | | Ref | | | Ref | | | Ref | | |
| Partially exposed | 0.882 | 0.532 | [0.596,1.306] | 1.227 | 0.305 | [0.830,1.813] | 0.880 | 0.523 | [0.594,1.304] | 1.227 | 0.306 | [0.829,1.816] |
| Heavily exposed | 0.547 | 0.003 | [0.366,0.819] | 1.765 | 0.004 | [1.202,2.592] | 0.550 | 0.004 | [0.367,0.824] | 1.761 | 0.004 | [1.196,2.591] |
| Post-exposure | 0.964 | 0.882 | [0.590,1.574] | 1.447 | 0.135 | [0.891,2.348] | 0.986 | 0.955 | [0.602,1.615] | 1.403 | 0.174 | [0.861,2.284] |
| Observations | 865 | | | 865 | | | 865 | | | 865 | | |

**Table S6: Measles1 Immunization at or before 9 months and between 10-12 months by Lockdown Exposure**
Odds ratios from logistic regressions of timeliness of Measles1 outcomes on indicators for exposure group, with unexposed children that turned 12 months old before March 2020 as the reference group. The sample is restricted to children with an immunization card and date of Measles1 recorded, if received. Adjusted regressions include indicators for survey month, low assets, and low caste as controls. OR = odds ratio

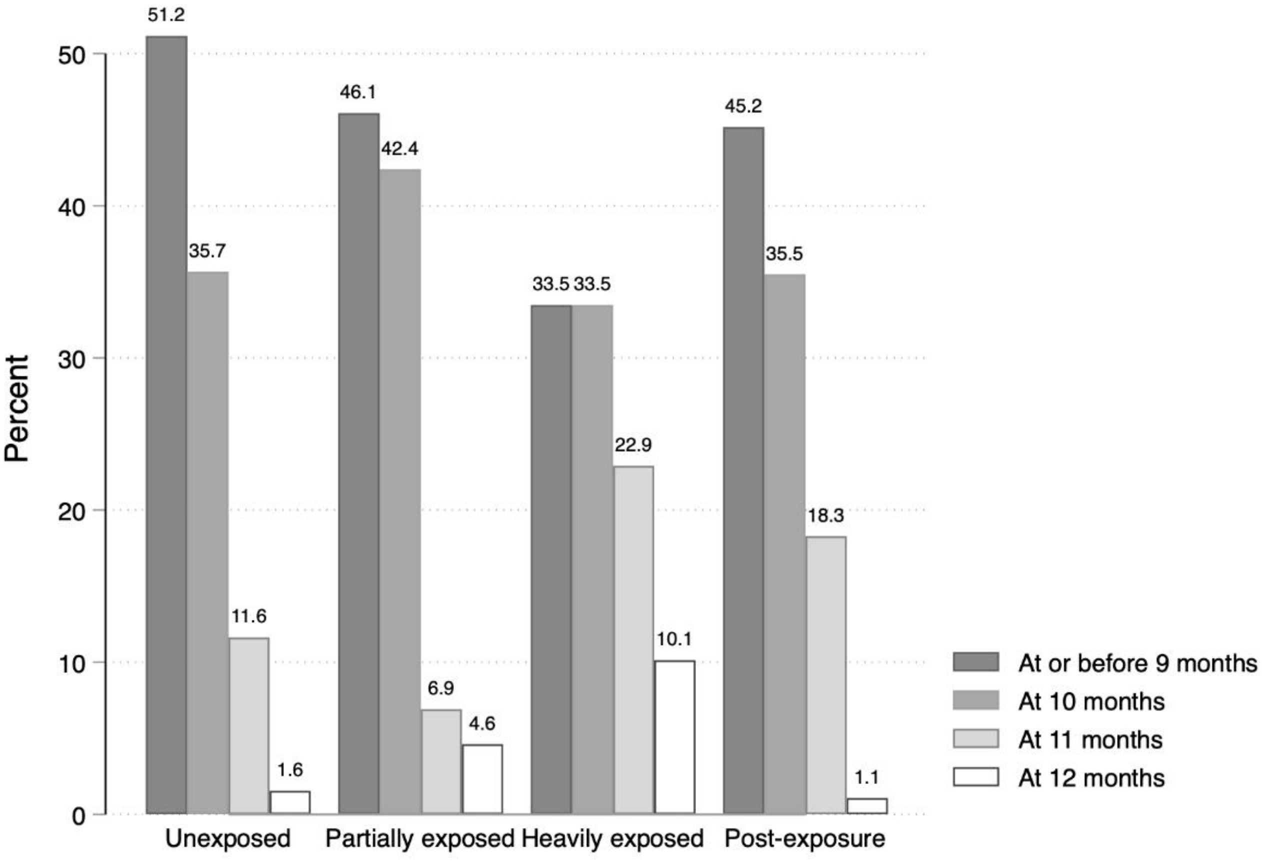

**Figure S3: Age at Measles1 Immunization by Lockdown Exposure**
The figure presents the breakdown of age at which children received their Measles1 immunization by lockdown exposure group, among children with any Measles1 on their immunization card.

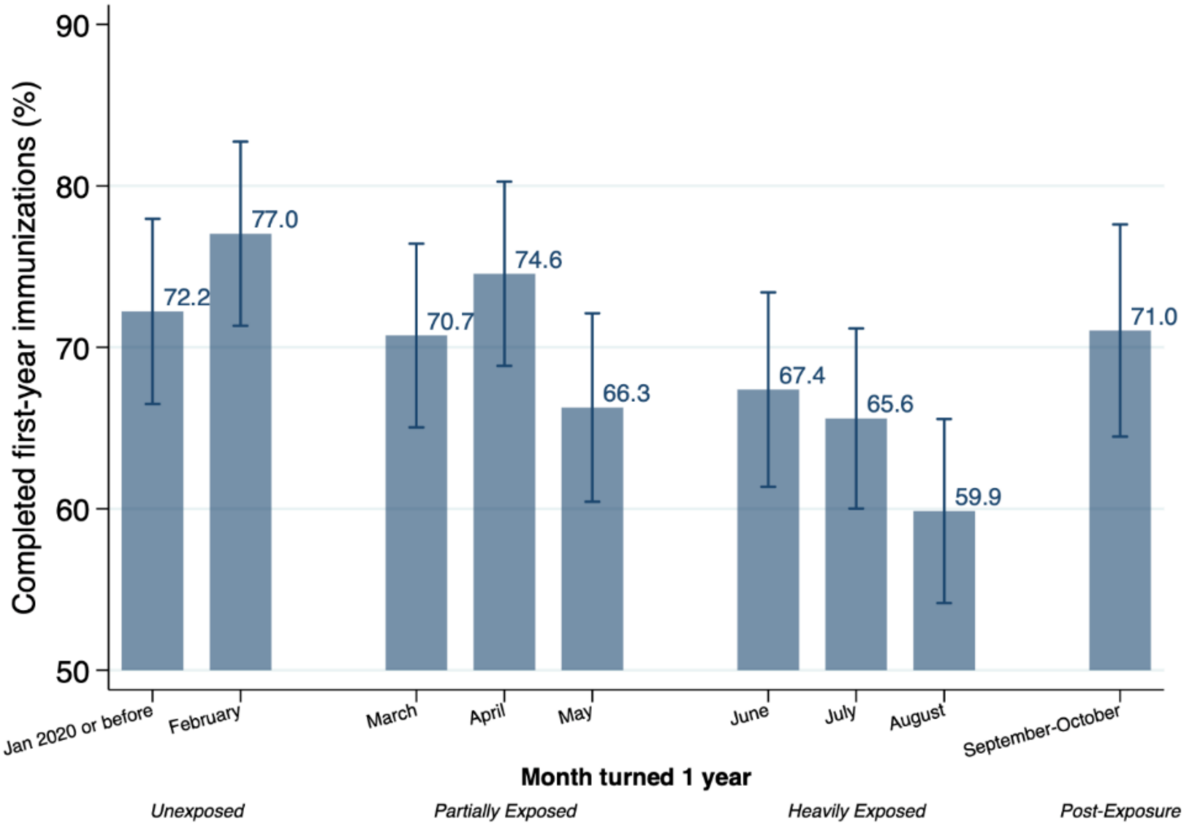

**Figure S4: Probability of Completed First-Year Immunizations at the time of Survey, by Cohort**
The figure presents unadjusted predicted probabilities of having received all key first-year immunizations by age cohort with 95% confidence intervals. The outcome is equal to one if the child received Pentavalent1, Pentavalent2, Pentavalent3, and Measles1 on the immunization card or at least 4 injectable immunizations per parent reports, if the immunization card was unavailable. Numbers underlying the figure are in table S7.

| | Observations | Probability | 95% Confidence Interval |
|---|---|---|---|
| Jan 2020 or before | 234 | 0.722 | [0.665,0.780] |
| February | 209 | 0.770 | [0.713,0.827] |
| March | 246 | 0.707 | [0.650,0.764] |
| April | 224 | 0.746 | [0.688,0.803] |
| May | 252 | 0.663 | [0.604,0.721] |
| June | 233 | 0.674 | [0.614,0.734] |
| July | 279 | 0.656 | [0.600,0.712] |
| August | 284 | 0.599 | [0.542,0.656] |
| September-October | 183 | 0.710 | [0.645,0.776] |
| Observations | 2,144 | | |

**Table S7: Probability of Completed First-Year Immunizations at the time of Survey, by Cohort**
The table presents the numbers underlying Figure S4.

**Panel A: Probability of Completed First-Year Immunizations**

| | Unadjusted | | | | Adjusted | | | |
|---|---|---|---|---|---|---|---|---|
| | From Immunization Card | | From Parent Reports | | From Immunization Card | | From Parent Reports | |
| | PR | 95% Confidence Interval | PR | 95% Confidence Interval | PR | 95% Confidence Interval | PR | 95% Confidence Interval |
| Unexposed | 0.841 | [0.797,0.884] | 0.644 | [0.578,0.710] | 0.840 | [0.796,0.883] | 0.644 | [0.579,0.709] |
| Partially exposed | 0.784 | [0.746,0.822] | 0.623 | [0.570,0.677] | 0.783 | [0.745,0.821] | 0.623 | [0.570,0.676] |
| Heavily exposed | 0.748 | [0.708,0.788] | 0.531 | [0.480,0.582] | 0.748 | [0.709,0.788] | 0.535 | [0.485,0.586] |
| Post-exposure | 0.786 | [0.710,0.862] | 0.610 | [0.501,0.719] | 0.791 | [0.715,0.867] | 0.590 | [0.478,0.702] |

**Panel B: Change in Completed First-Year Immunizations Relative to Unexposed Children**

| | Unadjusted | | | | | | Adjusted | | | | | |
|---|---|---|---|---|---|---|---|---|---|---|---|---|
| | From Immunization Card | | | From Parent Reports | | | From Immunization Card | | | From Parent Reports | | |
| | OR | p-value | 95% Confidence Interval | OR | p-value | 95% Confidence Interval | OR | p-value | 95% Confidence Interval | OR | p-value | 95% Confidence Interval |
| Unexposed | Ref | | | Ref | | | Ref | | | Ref | | |
| Partially exposed | 0.689 | 0.065 | [0.463,1.024] | 0.917 | 0.643 | [0.635,1.323] | 0.687 | 0.065 | [0.461,1.023] | 0.911 | 0.623 | [0.628,1.321] |
| Heavily exposed | 0.562 | 0.004 | [0.381,0.828] | 0.627 | 0.010 | [0.441,0.892] | 0.564 | 0.004 | [0.382,0.833] | 0.630 | 0.011 | [0.441,0.901] |
| Post-exposure | 0.695 | 0.199 | [0.398,1.212] | 0.868 | 0.607 | [0.505,1.490] | 0.721 | 0.261 | [0.407,1.275] | 0.791 | 0.412 | [0.451,1.386] |
| Observations | 1,287 | | | 966 | | | 1,287 | | | 966 | | |

**Table S8: Completed First-Year Immunizations from Immunization Card and Parent Reports by Lockdown Exposure**

In the main analysis we combined children with key first-year immunization status obtained from the immunization card and from parent reports. The table reports separate estimates of first-year immunization status separately for each data source. Panel A presents the predicted probabilities of having received all key first-year immunizations by lockdown exposure group and data source. The outcome is equal to one if the child received Pentavalent1, Pentavalent2, Pentavalent3, and Measles1 on the immunization card or at least 4 injectable immunizations per parent reports, if the immunization card was unavailable. Panel B presents odds ratios with 95% confidence intervals of the same outcome on indicators for exposure group, with unexposed children that turned 12 months old before March 2020 as the reference group. Adjusted regressions include indicators for survey month, low assets, and low caste as controls. PR = probability OR = odds ratio

| | Unadjusted | | | | | | Adjusted | | | | | |
|---|---|---|---|---|---|---|---|---|---|---|---|---|
| | Pentavalent1 | | | Pentavalent2 | | | Pentavalent1 | | | Pentavalent2 | | |
| | OR | p-value | 95% Confidence Interval | OR | p-value | 95% Confidence Interval | OR | p-value | 95% Confidence Interval | OR | p-value | 95% Confidence Interval |
| Unexposed | Ref | | | Ref | | | Ref | | | Ref | | |
| Partially exposed | 0.815 | 0.558 | [0.411,1.615] | 0.753 | 0.411 | [0.383,1.481] | 0.832 | 0.599 | [0.418,1.653] | 0.750 | 0.407 | [0.380,1.480] |
| Heavily exposed | 0.780 | 0.472 | [0.397,1.534] | 0.725 | 0.346 | [0.371,1.415] | 0.801 | 0.521 | [0.406,1.580] | 0.728 | 0.355 | [0.372,1.427] |
| Post-exposure | 1.082 | 0.883 | [0.377,3.111] | 0.658 | 0.366 | [0.265,1.633] | 1.001 | 0.999 | [0.339,2.956] | 0.642 | 0.354 | [0.252,1.639] |
| Observations | 1,287 | | | 1,287 | | | 1,287 | | | 1,287 | | |

**Table S9: Difference in Early Immunizations by Lockdown Exposure**

Odds ratios with 95% confidence intervals from logistic regressions of binary measures for receipt of key first-9 months immunizations on indicators for exposure group. Unexposed children that turned 12 months old before March 2020 are the reference group. The outcomes are respectively equal to one if the child has received BCG, Pentavalent1, and Pentavalent2 on the immunization card. The sample is restricted to children whose outcomes were measured from the immunization card. Adjusted regressions include indicators for survey month, low assets, and low caste as controls. OR = odds ratio

| | (1) | (2) | (3) | (4) | (5) | (6) | (7) |
|---|---|---|---|---|---|---|---|
| | Probability of Completed First-Year Immunizations (%) | | | PP Difference Heavily Exposed - Unexposed (95% CI) | p-value | PP Subgroup Difference (95% CI) | p-value |
| | Unexposed | Heavily Exposed | Post-Exposure | | | | |
| **Female** | 73.4 | 67.5 | 75.3 | -5.8 (-13.2 to 1.6) | 0.123 | | |
| **Male** | 75.1 | 61.8 | 66.9 | -13.4 (-20.5 to -6.3) | 0.000 | -7.5 (-17.7 to 2.8) | 0.152 |
| **High assets** | 77.1 | 70.4 | 74.1 | -6.5 (-13.3 to 0.3) | 0.061 | | |
| **Low assets** | 71.5 | 59.3 | 67.0 | -12.4 (-19.9 to -4.8) | 0.001 | -5.6 (-15.8 to 4.6) | 0.284 |
| **High parent education** | 79.0 | 73.2 | 67.8 | -5.8 (-12.6 to 1.1) | 0.101 | | |
| **Low parent education** | 70.1 | 57.1 | 72.7 | -13.0 (-20.3 to -5.7) | 0.000 | -7.2 (-17.3 to 2.8) | 0.160 |
| **High caste** | 74.8 | 66.8 | 69.2 | -8.0 (-13.9 to -2.2) | 0.007 | | |
| **Low caste** | 72.6 | 58.9 | 75.5 | -13.5 (-23.9 to -3.0) | 0.011 | -5.6 (-17.7 to 6.4) | 0.359 |
| **COVID other zones** | 73.0 | 65.3 | 72.7 | -7.7 (-13.4 to -2.0) | 0.009 | | |
| **COVID red zones** | 79.9 | 62.0 | 62.0 | -17.9 (-29.0 to -6.8) | 0.002 | -10.2 (-22.7 to 2.3) | 0.109 |

**Table S10: Changes in Completed Key First-Year Immunization Status Across Exposure Groups by Subgroup**
Columns 1-3 present the predicted probabilities of having received all key first-year immunizations for each exposure group by subgroup. Column 4 presents the percentage point difference in probability between the heavily exposed and unexposed groups by subgroup and Column 5 presents the p-value for this comparison. Column 6 presents the percentage point difference in subgroup differences and Column 7 presents the p-value for this comparison. 95% Confidence Intervals are included in parenthesis. Estimates include controls for survey month and whether the child had an immunization card.
